# Epidemiological and clinical characteristics in patients with SARS-CoV-2 antibody negative probable COVID-19 in Wuhan

**DOI:** 10.1101/2020.06.18.20134619

**Authors:** Limin Duan, Shuai Zhang, Mengfei Guo, E Zhou, Jinshuo Fan, Xuan Wang, Ling Wang, Feng Wu, Yang Jin

## Abstract

**Background:** Patients with suspected COVID-19 might be admitted to hospital. We aimed to describe the characteristic of SARS-CoV-2 antibody negative probable COVID-19 patients and give some suggestions to manage suspected COVID-19 patients.

**Methods:** We analyzed 616 confirmed COVID-19 patients and 35 SARS-CoV-2 antibody negative probable COVID-19 patients who were admitted in Wuhan Union Hospital from February 13, 2020 to February 16, 2020. Telephone interviews were conducted and medical records were reviewed for epidemiological, clinical, laboratory and radiographic data.

**Results:** Of the 35 SARS-CoV-2 antibody negative probable COVID-19 patients, all of them had tested at least 3 times of nucleic acid, 3 were believed to be non-SARS-CoV-2 infection. Compared with confirmed patients, antibody negative probable patients were younger (P=0.017), exhibited similar symptoms and chest CT images, had higher lymphocyte count (P=0.004) and albumin level (P<0.001), showed lower lactate dehydrogenase level (P=0.011) and erythrocyte sedimentation rate (P<0.001). During hospitalization, all the 35 patients had contacted with confirmed COVID-19 patients, but all used general face mask for protection and maintained a social distance of more than one meter from each other. All the isolation wards were kept ventilation and disinfected once a day. After discharged from hospital, all of them had negative nucleic acid tests and no one developed symptoms again.

**Conclusions:** The conditions of patients with AbN probable COVID-19 were less critical than those of patients with confirmed COVID-19. Room ventilation and daily disinfection, wearing face masks, and maintaining social distance might be helpful to prevent patients from hospital acquired COVID-19 infection.

## Introduction

Since December 2019, an ongoing outbreak of Coronavirus disease 2019 (COVID-19) has struck Wuhan, China and then spread all around the world.^1-3^ As of May 25, 2020, there have been a total of 5304772 laboratory-confirmed cases of COVID-19 reported to WHO globally, with 342029 world-wide ^4^. Currently, the early diagnosis of COVID-19 relies on specialized nucleic acid-based reverse transcription polymerase chain reaction (RT-PCR) testing. However, it has been reported that the positive rate of RT-PCR detection of nasopharyngeal swab and oropharyngeal was quite low.^5^ *Chen* et al. have listed several reasons why patients with suspected and confirmed COVID-19 should be hospitalized to hospital.^6^ Meanwhlie, World Health Organization (WHO) also suggested to screen and isolate all patients with suspected COVID-19 at the first point of contact with the health care system.^7^ Therefore, during the pandemic of COVID-19, many patients with suspected COVID-19 might be admitted to hospital, even some of them might be conventional infection.

Many studies have reported the epidemiology and clinical characteristics of patients with confirmed COVID-19.^1-3^ However, little was known about patients with suspected COVID-19. We investigated 133 suspected COVID-19 patients who were admitted to Wuhan Union Hospital from February 13, 2020 to February 16, 2020. Of the 133 patients, 35 patients had no laboratory evidence of COVID-19 infection even many SARS-CoV-2 nucleic acid and antibody tests were done. In this study, we aimed to compare the demographic, clinical, laboratory, and radiological characteristics of patients with confirmed COVID-19 and antibody negative (AbN) probable COVID-19. And we want to offer some suggestions to managing AbN probable COVID-19 patients since some of them may not be COVID-19 infection.

## Methods

### Study design and participants

This single-centre, retrospectivestudy (clinical trial identifier ChiCTR2000029770) was conducted at Wuhan Union Hospital (including two designated branch hospitals to treat patients with COVID-19). The study was approved by the Institutional Ethics Committee of Union Hospital, Tongji Medical College, Huazhong University of Science and Technology (20200036), the requirement for informed consent was exempted by the Ethics Committee.

The inclusion criteria were adult patients with confirmed and suspected COVID-19 on admission. Patients with negative RT-PCR testing results but didn’t test SARS-CoV-2 antibody were excluded. From February 13 to February 16, 2020, 616 confirmed COVID-19 patients and 133 suspected COVID-19 patients were admitted in Wuhan Union Hospital. Of the 133 suspected patients, 98 didn’t test SARS-CoV-2 antibody, 35 showed negative serological SARS-CoV-2 antibody results (Figure 1).

**Figure 1.**
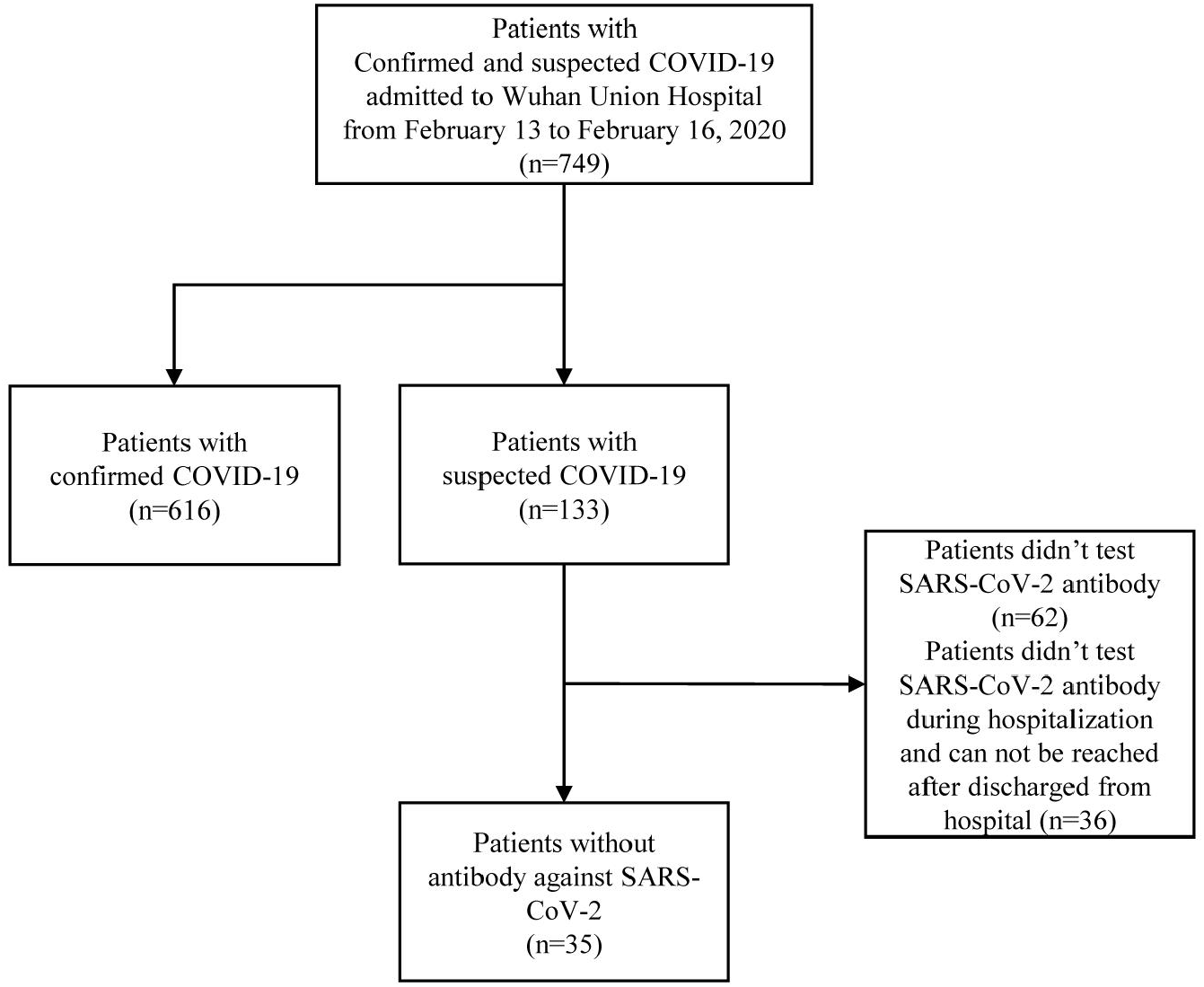
Study flow.

### Diagnosis of COVID-19

The diagnosis was made and clinical classification of COVID-19 was performed for the patients according to the Guidelines of the Diagnosis and Treatment of New Coronavirus Pneumonia (version 7) published by the National Health Commission of China.^8^ The diagnosis was established on the basis of (1) epidemiological history, (2) fever and/or other respiratory symptoms, (3) presence or absence of the imaging findings of novel coronavirus pneumonia, and (4) normal/reduced white blood cell count and normal/reduced lymphocyte count. A patient was taken as suspected case if she or he had epidemiological history, two of the aforementioned clinical presentations, or had no clear epidemiological history but had three of the clinical presentations. Suspected cases were considered to be confirmed if (1) real-time fluorescence RT-PCR for SARS-CoV-2 nucleic acid yielded positive results, or (2) Positive SARS-CoV-2 specific IgM and IgG antibody, or (3) Demonstration of a four-fold or greater increase in IgG antibody titer or conversion from a negative to a positive result between acute- and convalescent-phase serum specimens.

### Data Collection

Medical records were also reviewed for detail demographic and clinical information. All discharged patients with suspected COVID-19 infection were interviewed by telephone to collect the updated SARS-CoV-2 nucleic acid and antibody results. AbN probable COVID-19 patients were further investigated by telephone interview to record their potential contact and exposure history before and after admitted to hospital.

### Statistical Analysis

Categorical variables were presented as frequency rates and percentages, and continuous variables were expressed as mean±standard deviation (SD) if they were normally distributed or median (interquartile range [IQR]) if they were not. Proportions for categorical variables were compared using the χ^2^ test or Fisher’s exact test. Means for continuous variables were compared using independent group *t* test when the data were normally distributed. Otherwise, the Wilcoxon rank-sum test was employed. Missing data have been mentioned in the relevant tables, and there was no other missing data, unless otherwise noted. All statistical analyses were performed by using SAS software package (version 9.4).

## Results

### Demographic and clinical features in patients with confirmed and AbN probable COVID-19

The demographic and clinical characteristics at admission for the 616 confirmed and the 35 AbN probable patients are listed in Table 1. The proportion of female subjects in the confirmed COVID-19 group was 57.5%, while in AbN probable COVID-19 group was 57.1%. The median age of AbN probable patients was 55 (IQR, 37.0-68.0) years, which was significantly younger than confirmed patients (64 [IQR, 53.0-70.0] years). The median duration from symptoms onset to admission in AbN probable patients was 8 (IQR, 5-14) days, while in confirmed patients was 13 (IQR, 7-20) days, 27 (77.1%) of the AbN probable patients were hospitalized within 14 days after symptoms onset. On admission, the most common presenting symptoms of the AbN suspected COVID-19 patients were fever (19 [54.3%]), cough (19 [54.3%]), and weakness (12 [34.3%]). 11 (31.4%) of the AbN probable COVID-19 patients had chronic diseases, including hypertension (9 [25.7%]), chronic cardiac disease (3 [8.6%]), diabetes (4 [11.4%]), and chronic renal disease (1 [2.9%]). No significant differences were seen in any symptoms or comorbidities between confirmed patients and AbN probable patients.

**Table 1.**
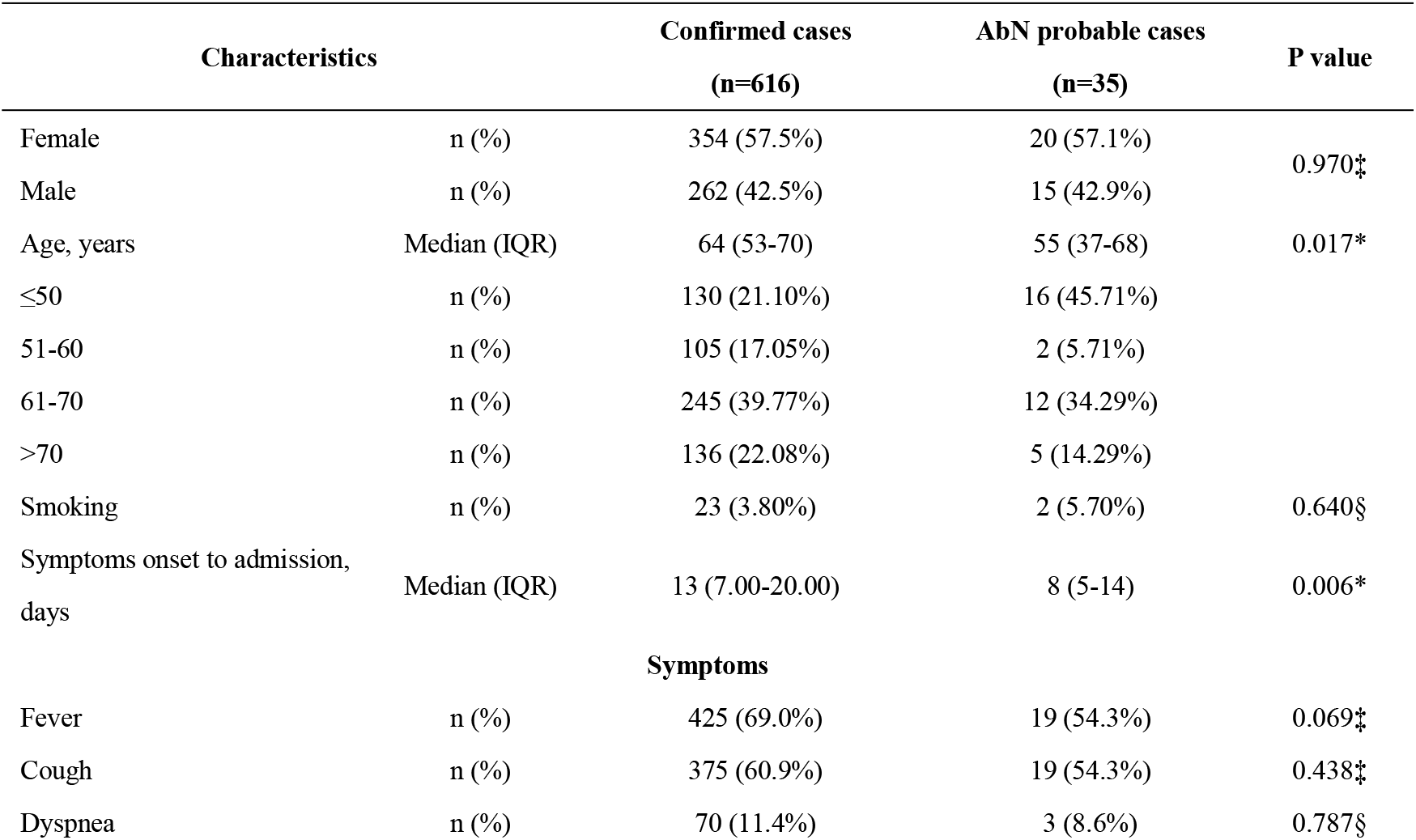
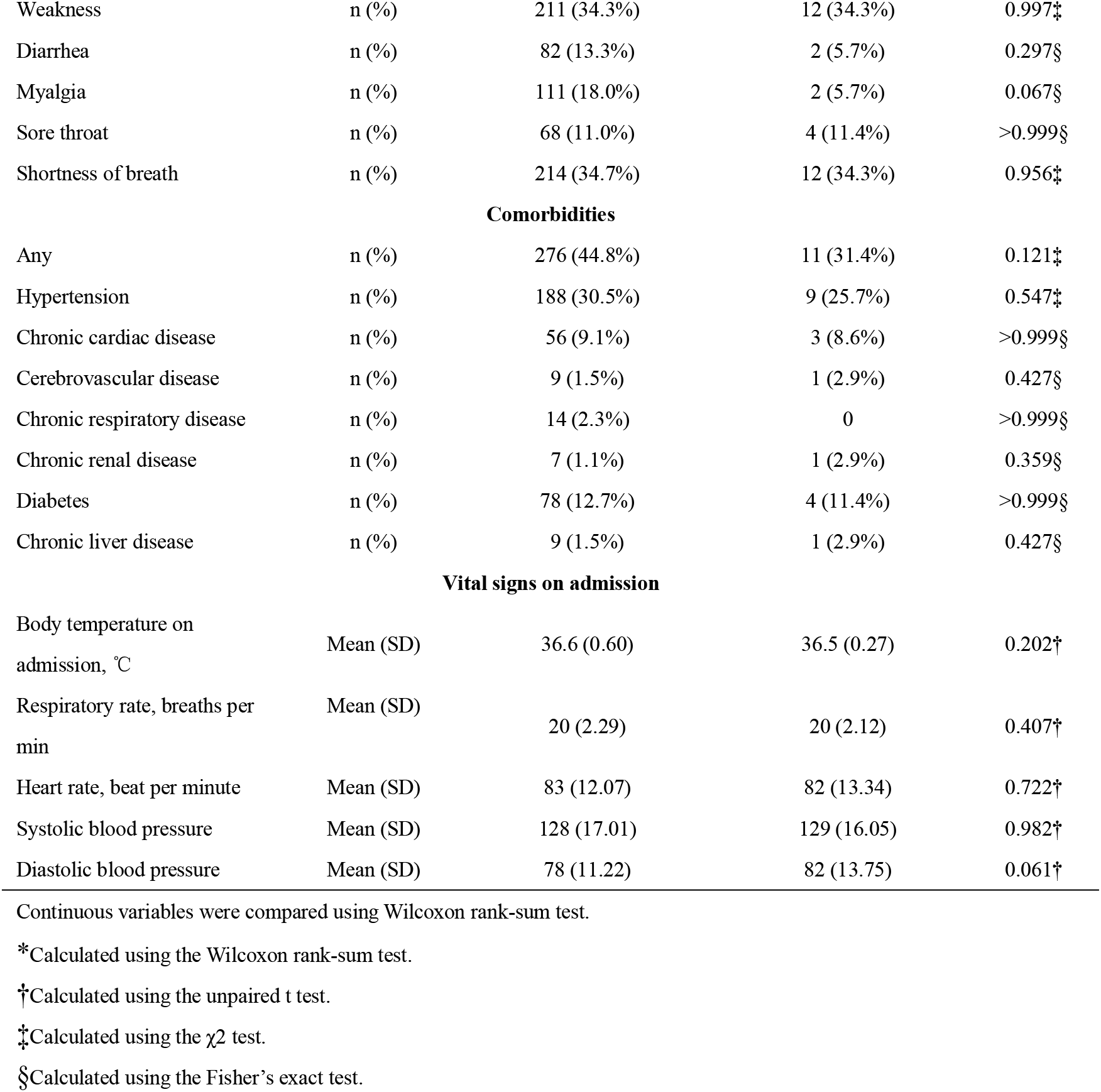
Demographic and Baseline Characteristics of patients with confirmed and AbN probable COVID-19.

### Laboratory and Radiological Findings in patients with confirmed and AbN probable COVID-19

Table 2 shows the laboratory findings and chest CT scan on admission. Compared with confirmed patients, AbN probable patients showed higher lymphocyte count (P=0.004), CD8 lymphocytes proportion (P=0.024) and albumin level (P<0.001). The lactate dehydrogenase (LDH, P=0.011), erythrocyte sedimentation rate (ESR, P<0.001) and fibrinogen level (P=0.001) were much lower in AbN probable patients than confirmed COVID-19 patients. Of the 35 AbN probable patients, 34 showed abnormal chest CT scan, 32 of them showed patchy or ground-glass opacity. During the course of the disease, 5 (14.3%) patients always showed unilateral lung involvement while 29 (82.9%) patients showed bilateral involvement. No significant differences were seen in chest CT scan between confirmed patients and AbN probable patients.

**Table 2.** Laboratory and Radiological Findings of patients with confirmed and AbN probable COVID-19.

| Characteristics | Confirmed cases<br>(n=616) | AbN probable cases<br>(n=35) | P value |
| --- | --- | --- | --- |
| **Laboratory Findings** |  |  |  |
| Leucocytes count, $\times 10^9/L$ | 5.66 (2.16) | 5.55 (1.64) | 0.756† |
| Neutrophils count, $\times 10^9/L$ | 3.80 (2.79) | 3.44 (1.36) | 0.454† |
| Lymphocyte count, $\times 10^9/L$ | 1.32(0.70) | 1.68 (0.62) | 0.004† |
| Hematocrit, % | 36.20 (33.50-39.50) | 38.80 (33.58-42.03) | 0.049* |
| Hemoglobin, g/L | 122.21 (16.12) | 128.09 (16.94) | 0.040† |
| Platelets count, $\times 10^9/L$ | 225.00 (172.00-293.25) | 212.50 (174.50-242.25) | 0.065* |
| Total bilirubin, $\mu\text{mol/L}$ | 13.80 (7.58) | 14.99 (5.81) | 0.366† |
| Direct bilirubin, $\mu\text{mol/L}$ | 4.31 (4.07) | 4.40 (2.35) | 0.907† |
| Albumin, g/L | 36.94 (4.86) | 41.44 (4.47) | <0.001† |
| Globin, g/L | 26.90 (24.50-30.00) | 23.70 (21.63-30.73) | 0.016* |
| ALT, U/L | 34.61 (35.85) | 27.12 (19.45) | 0.228† |
| AST, U/L | 31.32 (22.76) | 30.26 (25.46) | 0.794† |
| Urea nitrogen, mmol/L | 4.29 (2.30) | 4.31 (1.26) | 0.960† |
| Creatinine, µmol/L | 74.35 (22.46) | 77.34 (16.49) | 0.445† |
| GGT, U/L | 36.91 (40.88) | 33.24 (56.86) | 0.619† |
| Creatine kinase, U/L | 63.00 (45.50-97.00) | 80.00 (62.50-156.00) | 0.002* |
| LDH, U/L | 221.58 (84.35) | 183.24 (85.82) | 0.011† |
| CRP, mg/L | 20.72 (33.19) | 13.51 (29.34) | 0.237† |
| BNP, pg/ml | 61.38 (264.32) n=355 | 62.98 (127.50) n=24 | 0.977† |
| hs-TnT, ng/L | 2.70 (1.50-6.15) n=517 | 2.30 (1.10-3.65) n=28 | 0.135* |
| ESR, mm/h | 37.00 (20.00-61.00) n=408 | 13.00 (6.25-27.50) n=24 | <0.001* |
| Procalcitonin, ng/ml | 0.06 (0.04-0.15) n=157 | 0.04 (0.04-0.08) n=17 | 0.120* |
| D-dimer, mg/L | 1.69 (6.46) n=468 | 0.77 (1.69) n=29 | 0.443† |
| PT, s | 13.49 (1.44) n=557 | 13.18 (1.09) n=33 | 0.230† |
| APTT, s | 36.50 (33.70-39.80) n=557 | 35.70(33.05-39.40) n=33 | 0.358* |
| INR | 1.05 (0.16) n=557 | 1.02 (0.11) n=33 | 0.248† |
| Fibrinogen, g/L | 4.43 (1.97) n=557 | 3.26 (1.19) n=33 | 0.001† |
| CD3+T cell, % | 73.91 (11.24) n=544 | 75.38 (10.69) n=32 | 0.471† |
| CD4+T cell, % | 47.08 (40.89-53.82) n=544 | 44.06 (39.56-47.99) n=32 | 0.063* |
| CD8+T cell, % | 23.62 (8.77) n=544 | 27.22 (8.41) n=32 | 0.024† |
| IL-2, pg/ml | 2.84 (0.83) n=529 | 2.80 (0.64) n=31 | 0.780† |
| IL-4, pg/ml | 2.40 (1.06) n=529 | 2.41 (0.97) n=31 | 0.981† |
| IL-6, pg/ml | 37.76 (80.54) n=529 | 42.50 (66.59) n=31 | 0.748† |
| IL-10, pg/ml | 4.36 (10.86) n=529 | 3.38 (1.32) n=31 | 0.617† |
| TNF-α, pg/ml | 3.10 (2.22-4.86) n=529 | 2.89 (2.01-4.19) n=31 | 0.537* |
| IFN-γ, pg/ml | 2.56 (2.59) n=529 | 2.46 (1.41) n=31 | 0.833† |
| **Radiological Findings** |  |  |  |
| Unilateral | 64 (10.40%) | 5 (14.70%) | 0.426‡ |
| Bilateral | 552 (89.60%) | 29 (85.30%) |  |
| Patchy and ground-glass opacity | 589 (95.60%) | 32 (94.10%) | 0.659§ |

Continuous variables were expressed as mean (SD) if they were normally distributed or median (IQR) if they were not.
Abbreviations: ALT, Alanine aminotransferase; AST, Aspartate aminotransferase; GGT, Gamma-Glutamyl Transferase; LDH, Lactate dehydrogenase; BNP, type B natriuretic peptide; hs-TnT1, High sensitive Troponin I; CRP, C-reactive protein; ESR, Erythrocyte sedimentation rate; PT, Prothrombin time; APTT, Activated partial thromboplastin time; INR, International normalized ratio; IL-2, Interleukin-2; IL-4, Interleukin-4; IL-6, Interleukin-6; IL-10, Interleukin-10; TNF-α, Tumor necrosis factor-α; IFN-γ, interferon-γ.
\*Calculated using the Wilcoxon rank-sum test.
†Calculated using the unpaired t test.
‡Calculated using the $\chi^2$ test.
§Calculated using the Fisher's exact test.

**Table 3.**
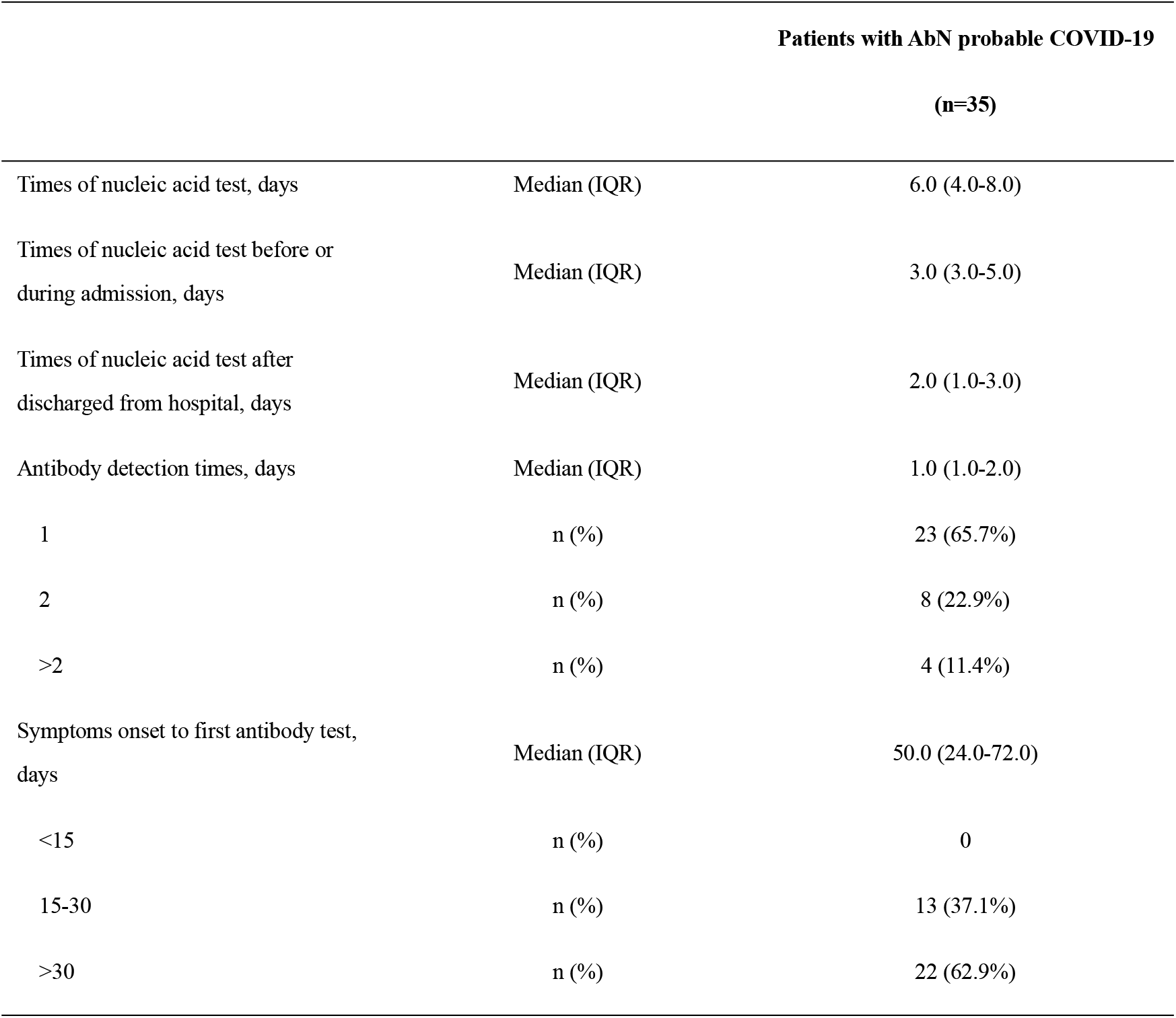
SARS-CoV-2 nucleic acid and antibody detection in patients with AbN probable COVID-19.

### Epidemiology and pathogen detection in patients with AbN probable COVID-19

All the AbN probable patients had tested at least 3 times of nucleic acid, the median times of SARS-CoV-2 nucleic acid test was 6.0 (IQR, 4.0-8.0). Among the 35 AbN probable COVID-19 patients, the duration from symptoms onset to first antibody test was 50.0 (IQR, 24.0-72.0) days, 12 patients had their antibody test two or more times, 8 patients tested the antibodies in two or more places in different time (Table 1). Figure 2 shows the time lines of SARS-CoV-2 nucleic acid and antibody test in the 12 patients who tested antibody two or more times. Another 23 AbN probable COVID-19 patients who tested antibody only once (Figure S1). During hospitalization, 20 of the AbN probable patients had tested serological mycoplasma and chlamydia antibodies, 15 of them showed positive mycoplasma or chlamydia IgG antibodies. Of the 35 AbN probable patients, a family cluster of 3 patients were admitted to the hospital and lived in one isolation ward. Table S1 shows the pathogens examined by the 3 patients. The 3 patients were eventually diagnosed as non-COVID-19 infection.

**Figure 2.**
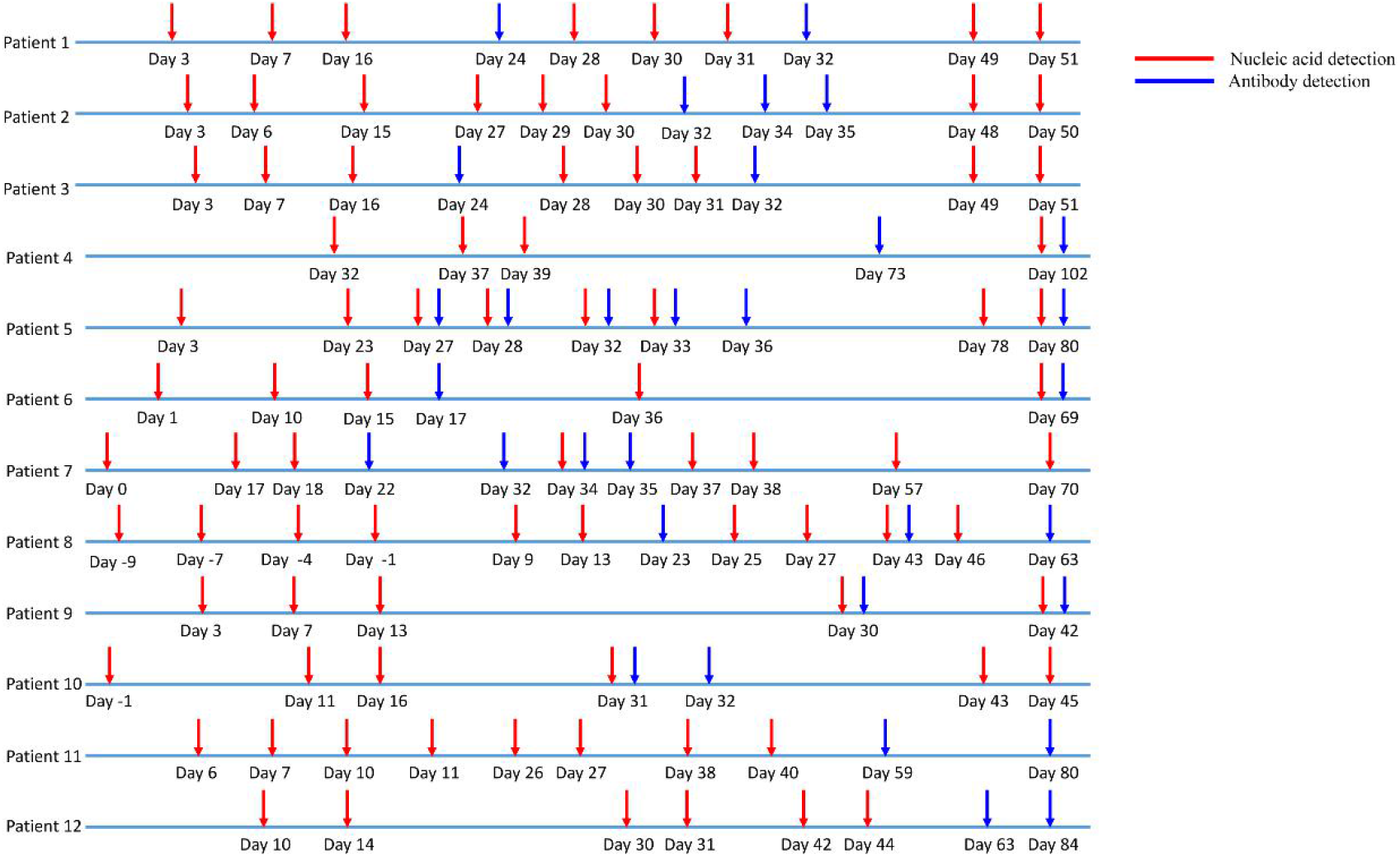
Timeline of nucleic acid and antibody detection of the 12 AbN probable COVID-19 patients who had tested SARS-CoV-2 more than once. Day 0 refers to the date of symptoms onset.

Of the 35 AbN probable COVID-19 patients, 14 had close contacted with confirmed or suspected COVID-19 patients before admission, 21 patients don’t know whether they had contacted patients with COVID-19. All the 35 patients had contacted with confirmed patients in hospital (in fever outpatient clinics or isolation wards), 32 patients shared the same room with confirmed COVID-19 patients. During hospitalization, the 35 patients almost wore face mask all the time, even when sleeping. All the patients except the 3 index family members claimed that they had maintained a distance of more than one meter when contacting with other patients. Additional measures were also taken by hospital and health-care workers to prevent patients from hospital acquired infection: 1) All health-care workers should disinfect their hands before contact with another patient; 2) Maintaining ventilation; 3) Surface disinfection of isolation wards every day; 4) Air disinfection for at least 1 hour per day. After discharged from hospital, all of them had negative SARS-CoV-2 nucleic acid tests and no one developed symptoms again.

## Discussion

Our study compares the clinical features between patients with confirmed COVID-19 and those with AbN probable COVID-19. We found that, compared with features in patients with confirmed COVID-19, patients with AbN probable COVID-19 were younger, showed higher lymphocyte count, albumin level and had lower LDH and ESR level. Previous studies have shown older age, lower lymphocyte count and higher levels of LDH to be associated with severe COVID-19 at admission.^9-10^ Another study demonstrated that decreased albumin was independent risk factor which influence the improvement of COVID-19 patients.^11^ Therefore, the conditions of patients with AbN probable COVID-19 were less critical than those patients with confirmed COVID-19. However, no significant differences were seen in any symptoms and chest CT findings between confirmed and AbN probable patients, suggesting that it’s hard for physicians to distinguish confirmed and AbN probable COVID-19 patients on admission.

Several reasons might contribute to the negative antibody test in the 35 patients: 1) false negative results of antibody test; 2) the delay of SARS-CoV-2 antibody production; 3) some of the them were conventional infection rather than COVID-19 infection. A study in our hospital showed the sensitivity of IgM and IgG test in suspected COVID-19 patients were 87.5% and 70.8%, respectively.^12^ A study in Italy even showed that IgG had 100% sensitivity on day 12 from symptoms onset.^13^ Two independent studies also demonstrated that on day 19 and day 15, the SARS-CoV-2 antibody could be tested on all the confirmed COVID-19 patients.^14^ In our study, a family cluster of 3 patients were diagnosed as non-COVID-19 infection. Of the other 32 AbN probable patients, 23 tested theSARS-CoV-2 antibody once, 9 patients had their antibody test two or more times, 8 patients even tested the SARS-CoV-2 antibodies in two or more places in different time. All of them had their antibody test after 15 days of symptoms onset. Therefore, based on the reported sensitivity and specificity of SARS-CoV-2 antibody tests, it is very likely that most of the AbN probable patients were not infected with COVID-19.

During the pandemic of COVID-19, early diagnosis and isolation of infections are essential for preventing further spread of COVID-19. WHO suggested to screen and isolate all patients with suspected COVID-19 at the first point of contact with the health care system.^7^ However, isolate all suspected patients might put some patients in danger, since some of them might not be infected with SARS-CoV-2. While conducting telephone interviews, we found those AbN probable or suspected patients have been suffering much anxiety and depress so far. A study of Vietnam showed suspected COVID-19 patients had a lower health-related quality of life and had a lower health-related quality of life.^15^ Under this circumstance, measures should be taken to protect suspected patients from hospital acquired COVID-19 infection. Of the 35 AbN probable cases, 32 had contacted and shared one room with confirmed COVID-19 patients. However, none of them showed any symptoms or positive SARS-CoV-2 nucleic acid results after discharged from hospital. These results demonstrated that none of the 35 AbN probable patients got hospital acquired COVID-19 infection. Since it is impossible for us to find exposure control for those patients, we can’t draw any conclusions about how to protect such patients. However, the following measures taken by hospital, patients and health-care workers might protect them from hospital acquired COVID-19 infection: 1) Daily disinfection isolation ward; 2) Maintaining ventilation in isolation wards; 3) Patients should wear surgical mask and keep at least 1 meter away from each other; 4) Health-care workers should disinfection their hands before touching patients. If possible, we also suggest to quarantine the confirmed and suspected COVID-19 patients separately.

Our study had several limitations. First, this is a retrospective study and there is no definite evidence that the AbN probable patients were infected with other pathogens. Second, not all suspected COVID-19 patients had tested SARS-CoV-2 antibody, those patients were excluded from our study and may cause selection bias. Third, no exposure control could be founded for the 35 AbN probable COVID-19 patients, we could not determine which measure are useful to protect patients from hospital acquired COVID-19 infection.

## Conclusions

Our study demonstrated that the conditions of patients with AbN probable COVID-19 were less critical than those of patients with confirmed COVID-19. But in fact, we cannot distinguish without SARS-CoV-2 infection from suspected COVID in clinical practice. We firstly points out the realistic dilemma about managing patients with suspected COVID-19: some patients without SARS-CoV-2 infection may be considered as suspected COVID-19 cases and share one isolation ward with confirmed COVID-19 patients in hospital. Wearing face mask, maintaining social distance, room ventilation and disinfection might be useful to protect patients from hospital acquired SARS-CoV-2 infection in the pandemic.

## Data Availability

We don't share the data of patients.

## Acknowledgments

We thank all the patients who consented to donate their data for analysis and the medical staff members who are on the front line of caring for patients.

## Funding

This work was supported by the State Project for Essential Drug Research and Development, China (no. 2019ZX09301001) and the National Natural Science Foundation of China (no. 81770096 and no. 81700091).The corresponding author had full access to all the data in the study and had final responsibility for the decision to submit for publication.

## Contributors

YJ and FW designed the study, LD, MG, EZ, JF, XW and LW collected the epidemiological and clinical data. LD, MG, and EZ summarized all data. SZ, LD and MG analyzed and drafted the manuscript. All authors revised the final manuscript.

## Declaration of interests

We declare no competing interests.

## Notes

### Competing Interest Statement

The authors have declared no competing interest.

## References

1. Wu F, Zhao S, Yu B, et al. A new coronavirus associated with human respiratory disease in China. Nature. 2020; published online Feb 3. https://doi:10.1038/s41586-020-2008-3.

2. Zhu N, Zhang D, Wang W, et al. A Novel Coronavirus from Patients with Pneumonia in China, 2019. The New England journal of medicine. 2020; 382(8):727–733.

3. Huang C, Wang Y, Li X, et al. Clinical features of patients infected with 2019 novel coronavirus in Wuhan, China. The Lancet. 2020; Feb 15; 395(10223):497–506.

4. World Health Organization. Novel coronavirus (COVID-19): situation report—126. https://www.who.int/docs/default-source/coronaviruse/situation-reports/20200525-covid-19-sitrep-126.pdf?sfvrsn=887dbd66_2 (Accessed May 26, 2020).

5. Rui L, Huan H, Fang L, et al. Positive rate of RT-PCR detection of SARS-CoV-2 infection in 4880 cases from one hospital in Wuhan, China, from Jan to Feb 2020. Clin Chim Acta. 2020; 505:172–175.

6. Chen S, Zhang Z, Yang J, et al. Fangcang shelter hospitals: a novel concept for responding to public health emergencies. Lancet. 2020; 395(10232):1305–1314.

7. WHO. Clinical management of severe acute respiratory infection when COVID-19 is suspected. https://www.who.int/publications-detail/clinical-management-of-severe-acute-respiratory-infection-when-novel-coronavirus-(ncov)-infection-is-suspected (Accessed Mach 13, 2020).

8. National Health Commission of China. The Guidelines of the Diagnosis and Treatment of New Coronavirus Pneumonia (version7). http://www.gov.cn/zhengce/zhengceku/2020-03/04/content_5486705.htm (Accessed March 6, 2020).

9. Jiao Gong, Jingyi Ou, Xueping Qiu, et.al. A Tool to Early Predict Severe Corona Virus Disease 2019 (COVID-19) A Multicenter Study Using the Risk Nomogram in Wuhan and Guangdong. China. Clin Infect Dis. 2020; published online Apr 16. https://doi:10.1093/cid/ciaa443.

10. Dong Ji, Dawei Zhang, Jing Xu. et.al. Prediction for Progression Risk in Patients With COVID-19 Pneumonia: The CALL Score. Clin Infect Dis. 2020; published online Apr 9. https://doi:10.1093/cid/ciaa414.

11. Zhang J, Wang X, Jia X, et.al. Risk factors for disease severity,unimprovement, and mortality of COVID-19 patients in Wuhan, China. Clinical Microbiology and Infection. 2020; https://doi.org/10.1016/j.cmi.2020.04.012.

12. Fei Xiang, Xiaorong Wang, Xinliang He, et.al. Antibody Detection and Dynamic Characteristics in Patients with COVID-19. Clin Infect Dis. 2020; published online Apr 19. https://doi:10.1093/cid/ciaa461.

13. Andrea Padoan, Chiara Cosma, Laura Sciacovelli, et al. Analytical Performances of a Chemiluminescence Immunoassay for SARS-CoV-2 IgM/IgG and Antibody Kinetics. Clin Chem Lab Med. 2020; published online Apr 16. https://doi:10.1515/cclm-2020-0443.

14. Juanjuan Zhao, Quan Yuan, Haiyan Wang, et al. Antibody Responses to SARS-CoV-2 in Patients of Novel Coronavirus Disease 2019. Clin Infect Dis. 2020; published online Mar 28. https://doi:10.1093/cid/ciaa344.

15. Hoang C Nguyen, Minh H Nguyen, Binh N Do, et al. People With Suspected COVID-19 Symptoms Were More Likely Depressed and Had Lower Health-Related Quality of Life: The Potential Benefit of Health Literacy. J Clin Med.2020, 9, 965.

